# Parasites and their protection against COVID-19- Ecology or Immunology?

**DOI:** 10.1101/2020.05.11.20098053

**Authors:** Kenneth Ssebambulidde, Ivan Segawa, Kelvin M. Abuga, Vivian Nakate, Anthony Kayiira, Jayne Ellis, Lillian Tugume, Agnes N. Kiragga, David B. Meya

## Abstract

**Background:** Despite the high infectivity of SARS-CoV-2, the incidence of COVID-19 in Africa has been slower than predicted. We aimed to investigate a possible association between parasitic infections and COVID-19.

**Methods:** An ecological study in which we analysed WHO data on COVID-19 cases in comparison to WHO data on helminths and malaria cases using correlation, regression, and Geographical Information Services analyses.

**Results:** Of the global 3.34 million COVID-19 cases and 238,628 deaths as at May 4^th^ 2020, Africa reported 0.029/3.3 million (0.88%) cases and 1,064/238,628 (0.45%) deaths. In 2018, Africa reported 213/229 million (93%) of all malaria cases, 204/229 million (89%) of schistosomiasis cases, and 271/1068 million (25%) of soil-transmitted helminth cases globally. In contrast, Europe reported 1.5/3.3 million (45%) of global COVID-19 cases and 142,667/238,628 (59%) deaths. Europe had 5.8/1068 million (0.55%) soil-transmitted helminths cases and no malaria/schistosomiasis cases in 2018. We found an inverse correlation between the incidence of COVID-19 and malaria (r −0.17, p =0.002) and COVID-19 and soil-transmitted helminths (r −0.25, p <0.001). Malaria-endemic countries were less likely to have COVID-19 (OR 0.51, 95% CI 0.29-0.90; p =0.02). Similarly, countries endemic for soil-transmitted helminths were less likely to have COVID-19 (OR 0.24, 95% CI 0.13-0.44; p <0.001), as were countries endemic for schistosomiasis (OR 0.22, 95% CI 0.11-0.45; p<0.001).

**Conclusions:** One plausible hypothesis for the comparatively low COVID-19 cases/deaths in parasite-endemic areas is immunomodulation induced by parasites. Studies to elucidate the relationship between parasitic infections and susceptibility to COVID-19 at an individual level are warranted.

## Background

Coronavirus disease 2019 (COVID-19) is caused by the severe acute respiratory syndrome coronavirus 2 (SARS-CoV-2). This disease started in China in December 2019. Globally, as at May 4^th^ 2020, there have been over 3.4 million confirmed COVID-19 cases and 238,628 deaths (1). There is, however, a notable disparity in the distribution of COVID-19 cases and deaths across the WHO regions with <1% of global cases reported in Africa to date where 17% of the global population lives. It may be conceivable that the distribution of the confirmed cases is highest in resource-rich settings due to availability of diagnostics alone, however, other mechanisms for the apparent lower incidence in Africa warrant investigation.

COVID-19’s disease spectrum spans from being asymptomatic, to severe respiratory distress requiring intensive care unit admission with respiratory support (2). The severity of COVID-19 is associated with an overwhelming immune response to SARS-CoV-2 marked by elevated pro-inflammatory cytokines including interleukin 6 (IL-6), IL-1β, IL-2, and tumour necrosis factor alpha (TNF-α) constituting the cytokine release syndrome (3). Additionally, high IL-6 has been associated with an increased risk of death in individuals with severe COVID-19 (4). Moreover, treatment of individuals with severe COVID-19 using monoclonal antibodies inhibiting IL-6 signalling have been associated with reduced intensive care stay and resolution of disease (5).

Parasites including *Plasmodium, Schistosome*, and various soil-transmitted helminths (STH) such as *Ascaris*, Hookworm, and *Trichuris* are common in Africa. Most of these parasites have a long life span in their infected human host who remains largely asymptomatic even though sterilising immunity is rarely achieved (6,7). Parasites are proficient immune modulators as they induce an immunotolerogenic state in infected individuals by striking a balance between pro-inflammatory and anti-inflammatory responses. Their immunomodulatory potential has been exploited through the use of parasitic excretory secretory products in the treatment of inflammatory conditions like multiple sclerosis and inflammatory bowel disease (8-10). Thus, it is plausible that parasitic infections could impact the incidence and clinical severity of COVID-19 in different WHO regions.

In this hypothesis-generating study, we aimed to investigate a possible association between endemic parasitic infections and COVID-19 cases and related fatality in the six WHO regions.

## Methods

### Study design

In this cross-sectional ecological study, we extracted quantitative data on COVID-19 confirmed cases and deaths from the WHO Situation Report-104 as of May 4^th^ 2020 (1). This data was compared to data collected from the 2019 world malaria report for malaria estimated cases and reported deaths in 2018, the 2018 schistosomiasis status data report for schistosomiasis cases, and the 2018 soil-transmitted helminths report for soil-transmitted helminths cases (11-13). We abstracted country specific and aggregated WHO regional data on COVID-19 cases and deaths. The world malaria report provided country specific and aggregated regional data on estimated incident malaria cases and reported deaths in 2018. The CDC Yellow book provided additional data on malaria endemic countries (14-16). The WHO repository on neglected tropical diseases provided data on the number of individuals who require helminth preventive chemotherapy as an indicator for the country-specific prevalence of and endemicity status for schistosomiasis and soil-transmitted helminths. Endemic areas as those in which a particular disease is prevalent. We performed an ecologic analysis based on our ability to access aggregated data as individual-level data were not available.

### Study procedures

Two investigators (KS and IS) captured data using an electronic case report form and entered it into Microsoft Excel 2013 (Microsoft Corporation, WA, USA). Abstracted variables were (1) country; (2) WHO region; (3) incidence of COVID-19 cases and deaths up until May 4^th^ 2020 by WHO region; (4) estimated incidence of, and deaths due to malaria, schistosomiasis and soil-transmitted helminths infections in 2018 by WHO region and (5) the parasite-specific endemicity status for each WHO region. Only countries (and territories) with COVID-19 cases reported by the WHO Situation Report-104 were included in the country specific analyses. We assigned non-endemic countries zero cases, and or deaths for malaria, schistosomiasis and soil-transmitted helminths.

### Geographical Information services analysis

To generate the maps for malaria, schistosomiasis, soil-transmitted helminth infections, and COVID-19 cases and deaths, we plotted the number of cases for each of the diseases at a national scale. Then geographic information services (GIS) tools were used to produce a continuous surface prevalence at 1 Å~ 1 km spatial resolution for visualization. The GIS analysis was conducted with AcrGIS desktop 10.0 (Redlands, CA: Environmental Systems Research Institute, Inc., 1999).

### Statistical analysis

Statistical analyses were conducted using STATA 15.1 (StataCorp, College Station, TX, USA). Medians and interquartile ranges of continuous variables were reported. Countries were classified as either endemic or non-endemic for malaria, schistosomiasis and soil-transmitted helminths, and either having <600 or ≥600 COVID-19 cases as at May 4^th^ 2020. Categorical data were expressed as proportions with corresponding percentages. Odds ratios were derived from univariate logistic regressions with COVID-19 (<600 or ≥600) cases as the dependent variable. Continuous data not normally distributed were log-transformed, and analysed with univariable linear regression models, as appropriate. Thereafter, a logistic regression model was done to establish the association between malaria, schistosomiasis, soil-transmitted helminths and COVID-19 cases and deaths. All analyses were considered statistically significant at P <0.05.

## Results

### WHO regional distribution of COVID-19 and parasitic infections

The six WHO regions analysed had 215 countries and territories including the unclassified international conveyance (Diamond Princess) territory. There were 3,349,786 COVID-19 cases and 238,628 deaths globally as of May 4^th^ 2020. The median number of COVID-19 cases was 592 (IQR 79-4,532) and COVID-19 deaths was 12 (IQR 2-97) per country/territory. In 2018, there were an estimated 228,709,000 malaria cases, 229,145,093 schistosomiasis cases, and 1,068,088,261 soil-transmitted helminth cases globally. Globally, 87/206 (42.23%) countries and territories were endemic for malaria, 52/160 (32.50%) were endemic for schistosomiasis, and 85/181 (46.96%) were endemic for soil-transmitted helminths.

The European region had the highest proportion, 1,518,895/3,349,786 (45.34%) of global COVID-19 cases and 142,667/238,628 (59.79%) of COVID-19 deaths. Europe had no malaria or schistosomiasis cases reported in 2018 and was thus categorised as a non-endemic region for malaria and schistosomiasis infections. Europe also had the lowest proportion, 5,856,201/1,068,088,261 (0.55%) of soil-transmitted helminth infections globally in 2018. The WHO region of Americas had 1,384,641/3,349,786 (41.34%) of the global COVID-19 cases and 78,409/238,628 (32.86%) of COVID-19 deaths; with 0.9/228 million (0.41%) of malaria cases, 1.6/229 million (0.71%) of schistosomiasis cases, and 57/1068 million (5.38%) of the soil-transmitted helminths cases globally in 2018. Africa had the smallest proportion, 29,438/3,349,486 (0.88%) of COVID-19 cases and 1,064/238,628 (0.45%) COVID-19 deaths globally, with the highest proportion, 213/228 million (93.13%) of malaria cases, and 204/229 million (89.23%) of schistosomiasis cases, and the second highest proportion, 271/1068 million (25.42%) of soil-transmitted helminths cases in 2018 (**Table 1**).

**Table 1:** WHO regional summaries for COVID-19 and parasitic infections.

| **WHO Region** | **COVID-19 cases**<br>**n (%)** | **COVID-19 deaths**<br>**n(%)** | **Malaria cases**<br>**n(%)** | **Malaria Deaths**<br>**n(%)** | **Schisto. Cases**<br>**n(%)** | **STH Cases**<br>**n(%)** |
| --- | --- | --- | --- | --- | --- | --- |
| **Africa** | 29,438(0.88) | 1,064(0.45) | 213,000,000(93.13) | 73,276(94.75) | 204,470,200(89.23) | 271,534,897(25.42) |
| **Americas** | 1,384,641(41.34) | 78,409(32.86) | 929,000(0.41) | 337(0.44) | 1,620,830(0.71) | 57,510,278(5.38) |
| **Eastern**<br>**Mediterranean** | 200,609(5.99) | 7,871(3.30) | 4,900,000(2.14) | 3,320(4.29) | 19,955,408(8.71) | 63,973,610(5.99) |
| **European** | 1,518,895(45.34) | 142,667(59.79) | 0*(0.00) | 0*(0.00) | 0*(0.00) | 5,856,201(0.55) |
| **Western Pacific** | 151,444 (4.52) | 6,229(2.61) | 1,980,000(0.87) | 243(0.31) | 3,073,935(1.34) | 86,118,794(8.06) |
| **South-East Asia** | 64,047(1.91) | 2,375(1.00) | 7,900,000(3.45) | 164(0.21) | 24,720(0.01) | 583,094,481(54.59) |
| **Global** | **3,349,786(100)** | **238,628(100)** | **228,709,000(100)** | **77,340(100)** | **229,145,093(100)** | **1,068,088,261(100)** |

\*Europe is a non-endemic region for malaria and schistosomiasis. COVID-19, coronavirus disease 2019; Schisto., Schistosomiasis; STH, soil-transmitted helminths.

### Geographical Information Systems findings

We found that the global distribution of malaria, schistosomiasis, and soil-transmitted helminth infections were inversely related to COVID-19 cases. There was a less distribution of COVID-19 cases and deaths in areas with a high distribution of malaria, schistosomiasis and soil-transmitted helminth infections. In contrast, areas with no or a low distribution of malaria, schistosomiasis and soil-transmitted helminth infections had a high distribution of COVID-19 cases/deaths (**Figure 1, Supplementary Figure 1)**.

**Figure 1:**
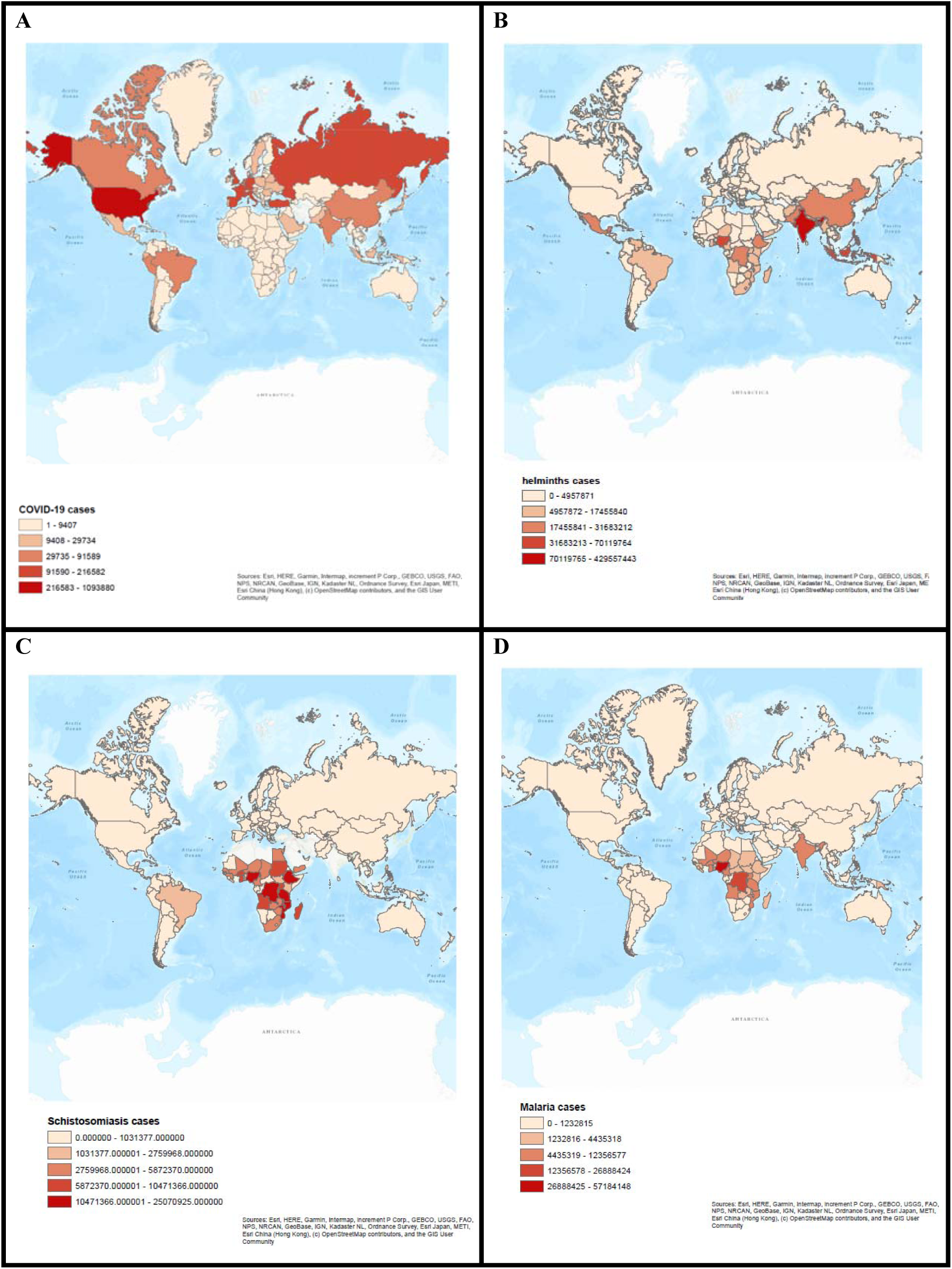
Geographical Information Services (GIS) analysis showing differential global distribution of COVID-19 cases (A), soil-transmitted helminth cases (B), schistosomiasis (C), and malaria (D).

### Association between COVID-19 and parasitic infections

The median number of COVID-19 cases was 315 (IQR 72-2,169) in malaria endemic countries and territories, 192 (IQR 58.2-705) in schistosomiasis endemic countries, and 270 (IQR 72-1,894) in soil-transmitted helminths endemic countries. Malaria, schistosomiasis and soil-transmitted helminth endemic countries had significantly lower median COVID-19 cases than their non-endemic counterparts (p <0.001). The median number of COVID-19 cases in the WHO African region was lower from those of Eastern Mediterranean (153 vs 2,344 cases, p <0.001) and Europe (153 vs 2,127 cases, p <0.001) (**Table 2**).

**Table 2:** Number of global COVID-19 cases in countries endemic and non-endemic for malaria, schistosomiasis and soil-transmitted helminths

|  | n (%) | COVID-19 cases<sup>‡</sup> | β coeff (95% CI) | P value<sup>¶</sup> |
| --- | --- | --- | --- | --- |
| Malaria |  |  |  |  |
| Non-endemic | 119 (57.8) | 1,136 (82, 7,755) | Reference |  |
| Endemic | 87 (42.2) | 315 (72, 2,169) | -0.81 (-1.58, -0.05) | 0.04 |
| Schistosomiasis |  |  |  |  |
| Non-endemic | 108 (67.5) | 1,655 (211, 12,322.5) | Reference |  |
| Endemic | 52 (32.5) | 192 (58.2, 705) | -1.70 (-2.56, -0.85) | <0.001 |
| STH |  |  |  |  |
| Non-Endemic | 96 (53.0) | 2,594 (531.5, 14855.5) | Reference |  |
| Endemic | 85 (47.0) | 270 (72, 1,894) | -1.87 (-2.60, -1.13) | <0.001 |
| WHO region |  |  |  |  |
| Africa | 48 (22.4) | 153 (42, 570) | Reference |  |
| E. Mediterranean | 22 (10.3) | 2,344 (592, 6193) | 2.29 (1.02, 3.55) | <0.001 |
| Europe | 61 (28.5) | 2,127 (795, 15558) | 2.86 (1.91, 3.81) | <0.001 |
| Americas | 54 (25.2) | 107 (16, 1229) | 0.22 (-0.76, 1.19) | 0.66 |
| South East Asia | 10 (4.7) | 612 (59, 8790) | 1.26 (-0.44, 2.97) | 0.15 |
| Western Pacific | 19 (8.9) | 144 (19, 8928) | 0.93 (-0.41, 2.26) | 0.17 |

<sup>‡</sup> Median cases and interquartile ranges.
<sup>¶</sup> P values were derived using univariate linear regression models. Coeff, coefficient; STH, soil-transmitted helminths.

There was a negative correlation between COVID-19 and malaria (r= −0.17, p =0.02), COVID-19 and soil-transmitted helminth infections (r= −0.25, p <0.001), and between COVID-19 and schistosomiasis (r= −0.38, p <0.001) **(Figure 2)**. Additionally, malaria endemic countries and territories were less likely to have 600 or more COVID-19 cases (OR 0.51, 95% CI 0.29-0.90; p =0.02). Similarly, schistosomiasis (OR 0.22, 95% CI 0.11-0.45; p <0.001) and soil-transmitted helminth (OR 0.24, 95% CI 0.13-0.44; p <0.001) endemic countries were less likely to have 600 or more COVID-19 cases. Africa was less likely than the Eastern Mediterranean or European region to have 600 or more COVID-19 cases using the univariate logistic regression model (**Table 3**). However, on adjusting for WHO region in multivariate analysis, there were no association between endemic status for malaria, schistosomiasis and soil-transmitted helminths and 600 or more COVID-19 cases (data not shown).

**Figure 2:**
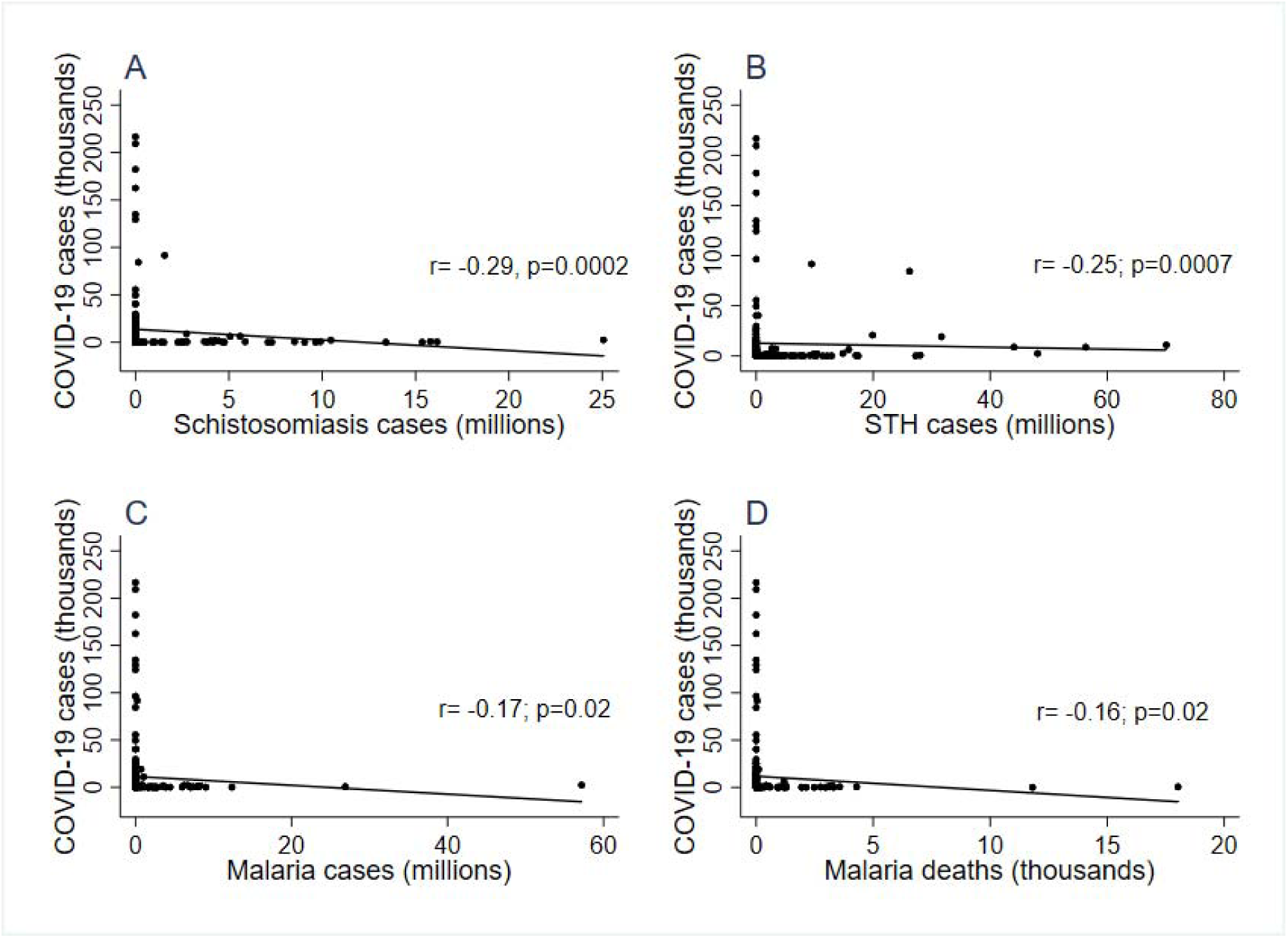
Scatter plots for COVID-19 cases and schistosomiasis cases (A), soil-transmitted helminths, STH (B), malaria cases (C) and malaria deaths (D).

**Table 3:** Associations between COVID-19 cases and endemicity of malaria, schistosomiasis and soil-transmitted helminths

|  | <600 COVID-19 cases, n (%) | ≥600 COVID-19 cases, n (%) | OR (95% CI) | P value |
| --- | --- | --- | --- | --- |
| Malaria |  |  |  |  |
| non-endemic | 50 (50.5) | 69 (65.7) | Reference |  |
| endemic | 51 (49.5) | 36 (34.3) | 0.51 (0.29, 0.90) | 0.02 |
| Schistosomiasis |  |  |  |  |
| Non-endemic | 36 (50.0) | 72 (81.8) | Reference |  |
| Endemic | 36 (50.0) | 16 (18.2) | 0.22 (0.11, 0.45) | <0.001 |
| STH |  |  |  |  |
| Non-endemic | 26 (33.3) | 70 (68.0) | Reference |  |
| Endemic | 52 (66.7) | 33 (32.0) | 0.24 (0.13, 0.44) | <0.001 |
| WHO region |  |  |  |  |
| Africa | 37 (33.9) | 11 (10.5) | Reference |  |
| E. Mediterranean | 6 (5.5) | 16 (15.2) | 8.97 (2.83, 28.5) | <0.001 |
| Europe | 14 (12.8) | 47 (44.8) | 11.29 (4.59, 27.76) | <0.001 |
| Americas | 36 (33.0) | 18 (17.1) | 1.68 (0.70, 4.05) | 0.25 |
| South East Asia | 5 (4.6) | 5 (4.8) | 3.36 (0.82, 13.78) | 0.09 |
| Western Pacific | 11 (10.1) | 8 (7.6) | 2.45 (0.79, 7.59) | 0.12 |

Odds ratios (OR) and P values were derived from univariate logistic regression models.
STH, soil-transmitted helminths; WHO, World Health Organization; E. Mediterranean, Eastern Mediterranean.

## Discussion

We found an inverse correlation between the number of COVID-19 cases and deaths, and the endemicity of parasitic infections globally. Africa had the highest burden of parasitic infections globally, and reported the lowest global proportion of COVID-19 confirmed cases/deaths of all WHO regions. In contrast, the parasitic-infection-non-endemic European region has the highest global proportion of COVID-19 cases/deaths. Our analyses showed that COVID-19 cases reduced with endemicity of malaria, schistosomiasis, or soil-transmitted helminth infections suggestive of a possible protective effect from COVID-19.

Nioni and Napoli (2020) reported that individuals in malaria endemic settings seem to be protected from COVID-19 (17). Their hypothesized ecological protection was based on molecular and genetic variations associated with malaria that make hosts less susceptible to COVID-19. We have now shown that together with malaria, schistosomiasis and soil-transmitted helminths may also confer some protection against COVID-19. Additionally, BCG vaccination has also been recently reported to correlate with low COVID-19 cases and deaths (18). This correlation was postulated to be related to non-specific immune stimulation by BCG that leads to protection against other microbes other than *Mycobacteria spps*. There is potentially cross protection from COVID-19 through yet-to-be discerned immune mechanisms.

Our findings have elicited more hypotheses behind this protection. Firstly, we hypothesise that the host immune response to parasitic infections entailing immunotolerance through induction of regulatory CD4^+^ T cells, and secretion of immunomodulatory cytokines (IL-10 and TGFβ), might be responsible for the protection from COVID-19 **(Figure 3)**. Moreover, severe COVID-19 has been reported to be related to an overly activated pro-inflammatory state that inhibition through IL-6 antagonism resulted in improvement of COVID-19 clinical severity (5,19). We, therefore, postulate that the lower proportion of COVID-19 confirmed clinical cases and deaths in Africa may be related to the immunomodulatory effects occasioned by parasitic infections endemic in this region. Of note, immune responses to parasitic infections in populations of individuals who relocate from areas of sustained transmission wane (20). Based on the differences in COVID-19 cases reported between endemic and non-endemic countries, we also suppose that parasite-induced immunomodulatory mechanisms wane among individuals who emigrate from parasite-endemic regions, with subsequent waning of population-based protection from COVID-19.

**Figure 3:**
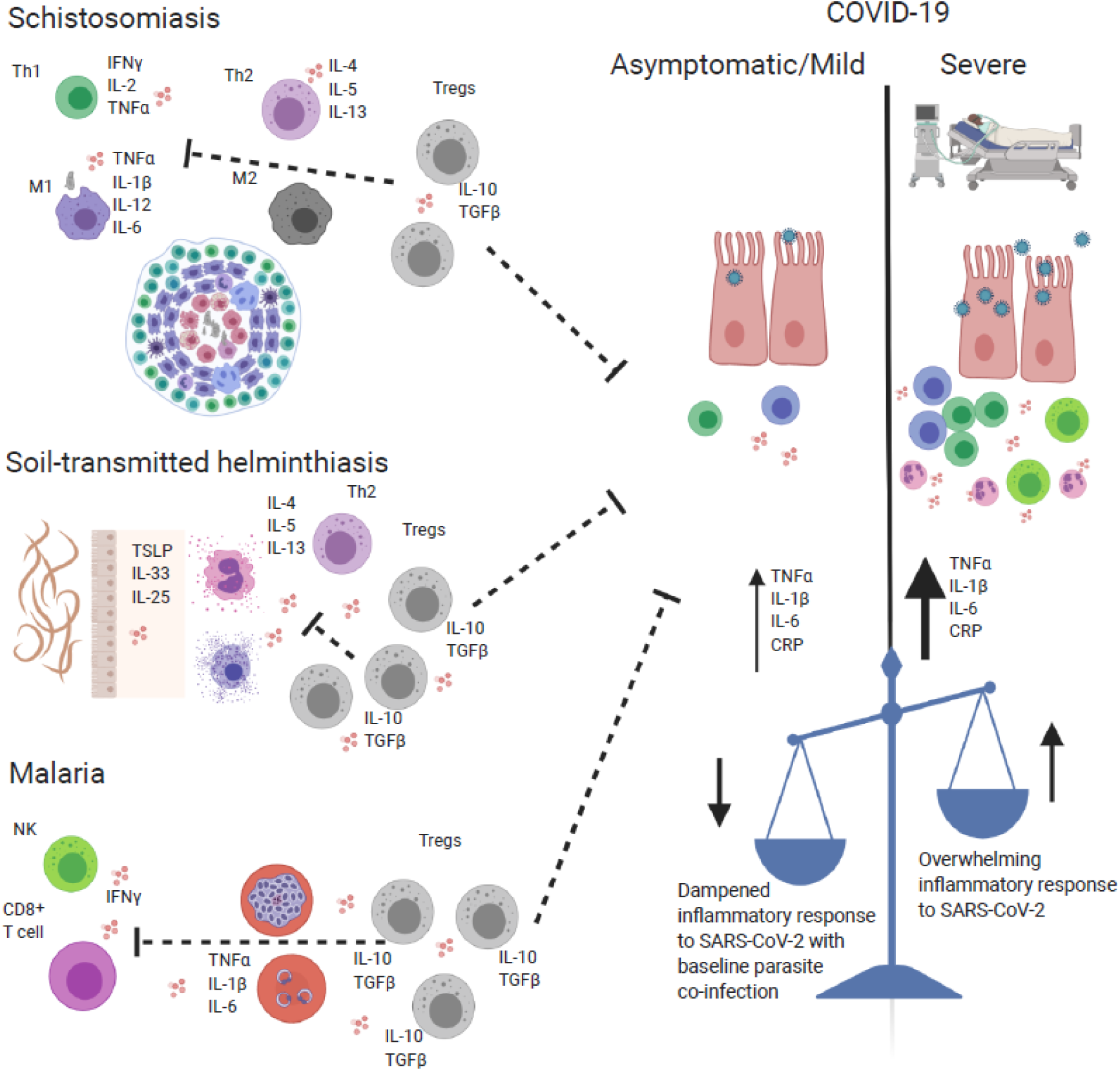
Immunoregulatory responses elicited by parasitic infections potentially dampen pro-inflammatory response to SARS-CoV-2 resulting into an asymptomatic or mild clinical COVID-19 presentation. Image created using resources at: www.biorender.com

Parasites have co-evolved with humans and thus far become proficient immune modulators. For instance, in schistosomiasis, there is a balance between type 1 and 2 immune response related cytokines (IFNγ, IL-4, IL-5, and IL-13) which are required to contain the *Schistosome* eggs in granulomas- and the immunoregulatory cytokines IL-10 which dampen these responses in order to limit immunopathology (21-24). Additionally, individuals in malaria endemic regions with stable transmission exhibit clinical immunity to malaria. This is a state in which individuals have detectable malaria parasitemia but remain clinically asymptomatic (25,26). The basis of clinical immunity to malaria has been attributed to immunoregulatory mechanisms including regulatory CD4^+^ T cells which secrete IL-10 and transforming growth factor beta (TGF-β) (27,28). These two cytokines are master immunomodulatory cytokines which block the effects of pro-inflammatory cytokines and thus induce an immunotolerant milieu. The immunotolerance marked by anti-inflammatory cytokines and regulatory T cells induced in individuals with parasitic infections arises from the need to limit pro-inflammatory-cytokine-related immunopathology. It also occurs for gut-dwelling helminths and thus cuts across parasitic infections (6). This immunotolerogenic state typical in individuals living in parasite-endemic areas has been previously reported to reduce vaccine efficacy in these settings in comparison to settings without a high burden of parasitic infections (29,30). For example, in Ethiopia, interferon gamma responses following BCG vaccination were noted to be lower in individuals with an active helminth infection. Furthermore, following anti-helminthic treatment, responses to BCG vaccination improved presumably due to clearance of immunomodulatory parasites (31). Parasite immunomodulatory mechanisms have been summarised in Figure 3.

However, the reasons underpinning the apparent low number of COVID-19 cases in settings with parasitic-infection endemicity is multifactorial. The low numbers could in part be related to limited testing due to limited resources dedicated to diagnostics. For example, data from Uganda shows 47,620 tests have been performed with 100 confirmed cases compared to data from the United Kingdom where 1,448,010 tests have been performed with 201,101 confirmed cases as at May 7^th^ 2020 (32,33). The low cases could also be related to limited cross-border transmission due to limited international travel (34). But also differences in climate, population pyramid, stage of the pandemic, and immunological differences due to cross protection by the endemic infections including but not limited to parasites (35).

## Strengths and limitations

Our study highlights a plausible immunologic hypothesis that parasitic infections could play an immune-protective role against SARS-CoV-2 infection or progression to a severe form of COVID-19 disease. However, we acknowledge a potential incongruence between regional/country and individual associations due to the ecologic bias associated with the study design, hence our findings are hypothesis generating only. Additionally, in this expanding pandemic, different countries and or continents are at different epidemiological phases of the current COVID-19 pandemic that the data we have used in our current analyses could change in the future. Secondly, we cannot rule out the possibility of misclassification of countries as high/low endemic settings as we utilised secondary data for our analysis. The potential non-differential misclassification bias would likely have produced an overestimation of the true odds ratio of the relationship between endemic countries and COVID-19 cases. Thirdly, there is a potential selection bias because we only included countries with COVID-19 cases reported by WHO in this analysis. Finally, our findings are limited by lack of data on confounding factors like population size, climate, immunogenetics/race, nutritional status, gender, other co-infections/co-morbidities, age stratification, and number of tests done per million people which need to be adjusted for and could potentially affect our observed correlations.

## Future Research

We recommend future research evaluating the immunological correlates of COVID-19 among individuals with parasitic co-infections to guide future interventions like SARS-CoV-2 vaccination in parasite endemic areas. We further recommend seroepidemiological studies to better characterise COVID-19 and parasite co-infection.

## Conclusion

We report a consistent inverse correlation between the incidence of COVID-19 and parasitic infections observed across WHO regions. These preliminary findings from an ecological analysis, support our hypothesis of a possible immune-modulatory mechanism induced by parasitic infections, which is protective against COVID-19 and warrants further investigation.

## Data Availability

Data is available on request

## Reference

1. WHO. Coronavirus Disease (COVID-19) Situtation report 104. World Heal Organ. 2020;(March 2020):1–7.

2. Verity R, Okell LC, Dorigatti I, Winskill P, Whittaker C, Imai N, et al. Estimates of the severity of coronavirus disease 2019: a model-based analysis. Lancet Infect Dis. 2020;3099(20):1–9.

3. Huang C, Wang Y, Li X, Ren L, Zhao J, Hu Y, et al. Clinical features of patients infected with 2019 novel coronavirus in Wuhan, China. Lancet. 2020;395(10223):497–506.

4. Ruan Q, Yang K, Wang W, Jiang L, Song J. Clinical predictors of mortality due to COVID-19 based on an analysis of data of 150 patients from Wuhan, China. Intensive Care Med. 2020;1–3.

5. Michot J-M, Albiges L, Chaput N, Saada V, Pommeret F, Griscelli F, et al. Tocilizumab, an anti-IL6 receptor antibody, to treat Covid-19-related respiratory failure: a case report. Ann Oncol. 2020;

6. Maizels RM, Balic A, Gomez-Escobar N, Nair M, Taylor MD, Allen JE. Helminth parasites - Masters of regulation. Immunol Rev. 2004;201:89–116.

7. Moreau E, Chauvin A. Immunity against helminths: Interactions with the host and the intercurrent infections. J Biomed Biotechnol. 2010;2010:428593.

8. Fleming JO, Isaak A, Lee JE, Luzzio CC, Carrithers MD, Cook TD, et al. Probiotic helminth administration in relapsing-remitting multiple sclerosis: A phase 1 study. Mult Scler J. 2011;17(6):743–54.

9. Benzel F, Erdur H, Kohler S, Frentsch M, Thiel A, Harms L, et al. Immune monitoring of Trichuris suis egg therapy in multiple sclerosis patients. J Helminthol. 2012;86(3):339–47.

10. Broadhurst MJ, Leung JM, Kashyap V, McCune JM, Mahadevan U, McKerrow JH, et al. IL- 22+ CD4+ T cells are associated with therapeutic Trichuris trichiura infection in an ulcerative colitis patient. Sci Transl Med. 2010;2(60).

11. WHO. World Malaria Report 2019. Geneva. World Health Organization Global Malaria Programme. 2019.

12. WHO. Schistosomiasis. Status of Schistosomiasis endemic countries [Internet]. 2018 [cited 2020 Apr 23]. Available from: https://apps.who.int/neglected_diseases/ntddata/sch/sch.html

13. WHO. Soil-transmitted helminthiases: Number of children (Pre_SAC and SAC) requiring preventive chemotherapy for soil-transmitted helminthiases. [Internet]. 2018 [cited 2020 Apr 23]. Available from: https://apps.who.int/neglected_diseases/ntddata/sth/sth.html

14. CDC. CDC 2019: Malaria Information and Prophylaxis, by country. Global Health, Division of Parasitic Diseases and Malaria [Internet]. 2019 [cited 2020 Apr 28]. Available from: https://www.cdc.gov/malaria/travelers/country_table/s.html

15. CDC. CDC Yellow Book 2014 Map: Health Information for International Travel. In: CDC Yellow Book 2014 Map. 2014.

16. CDC. CDC Yellow Book 2020: Health Information for International Travel. In 2020.

17. Nioi M, Napoli PE. Global Spread of Coronavirus Disease 2019 and Malaria: An Epidemiological Paradox. J Clin Med. 2020;9:1138.

18. Miller A, Reandelar J, Fasciglione K, Roumenova V, Li Y, Otazu HG. Correlation between universal BCG vaccination policy and reduced morbidity and mortality for COVID-19: an epidemiological study. medRxiv. 2020;6.

19. Zhang C, Wu Z, Li J-W, Zhao H, Wang G-Q. The cytokine release syndrome (CRS) of severe COVID-19 and Interleukin-6 receptor (IL-6R) antagonist Tocilizumab may be the key to reduce the mortality. Int J Antimicrob Agents. 2020 Mar; 105954.

20. Ghani AC, Sutherland CJ, Riley EM, Drakeley CJ, Griffin JT, Gosling RD, et al. Loss of population levels of immunity to malaria as a result of exposure-reducing interventions: Consequences for interpretation of disease trends. PLoS One. 2009;4(2).

21. Jenkins SJ, Hewitson JP, Jenkins GR, Mountford AP. Modulation of the host’s immune response by schistosome larvae. Parasite Immunol. 2005;27(10–11):385-93.

22. McManus DP, Bergquist R, Cai P, Ranasinghe S, Tebeje BM YH. Schistosomiasis — from immunopathology to vaccines. Semin Immunopathol. 2020;10(1007).

23. Butterworth AE. Immunological aspects of human schistosomiasis. Br Med Bull. 1998;54(2):357–68.

24. Caldas IR, Campi-Azevedo AC, Oliveira LFA, Silveira AMS, Oliveira RC, Gazzinelli G. Human schistosomiasis mansoni: Immune responses during acute and chronic phases of the infection. Acta Trop. 2008;108(2–3):109-17.

25. WHO. Malaria [Internet]. 2020 [cited 2020 Apr 24]. Available from: https://www.who.int/news-room/fact-sheets/detail/malaria

26. Doolan DL, Dobaño C, Baird JK. Acquired immunity to malaria. Clin Microbiol Rev. 2009 Jan;22(1):13–36.

27. Omer FM, Kurtzhals JA, Riley EM. Maintaining the immunological balance in parasitic infections: a role for TGF-beta? Parasitol Today. 2000 Jan;16(1):18–23.

28. Kumar R, Ng S, Engwerda C. The role of IL-10 in malaria: A double edged sword. Front Immunol. 2019;10(FEB):1–10.

29. Malhotra I, Mungai P, Wamachi A, Kioko J, Ouma JH, Kazura JW, et al. Helminth- and Bacillus Calmette-Guérin-induced immunity in children sensitized in utero to filariasis and schistosomiasis. J Immunol. 1999;162(11):6843–8.

30. Cooper PJ, Chico M, Sandoval C, Espinel I, Guevara A, Levine MM, et al. Human infection with Ascaris lumbricoides is associated with suppression of the interleukin-2 response to recombinant cholera toxin B subunit following vaccination with the live oral cholera vaccine CVD 103-HgR. Infect Immun. 2001;69(3):1574–80.

31. Elias D, Wolday D, Akuffo H, Petros B, Bronner U, Britton S. Effect of deworming on human T cell responses to mycobacterial antigens in helminth-exposed individuals before and after bacille Calmette-Guérin (BCG) vaccination. Clin Exp Immunol. 2001;123(2):219–25.

32. Moh U. Coronavirus (Pandemic) COVID-19 [Internet]. 2020 [cited 2020 May 7]. Available from: https://www.health.go.ug/covid/

33. GOV.UK. Guidance Number of coronavirus (COVID-19) cases and risk in the UK [Internet]. 2020 [cited 2020 May 7]. Available from: https://www.gov.uk/guidance/coronavirus-covid-19-information-for-the-public

34. Haider N, Yavlinsky A, Simons D, Osman AY, Ntoumi F, Zumla A, et al. Passengers’ destinations from China: Low risk of Novel Coronavirus (2019-nCoV) transmission into Africa and South America. Epidemiol Infect. 2020;363(December 2019).

35. Hopman J, Allegranzi B, Mehtar S. Managing COVID-19 in Low- and Middle-Income Countries. JAMA. 2020 Apr 28;323(16):1549–50.

